# Quantifying the impact of UVC in reducing airborne pathogen transmission and improving energy efficiency for healthy buildings: Kahn-Mariita equivalent ventilation model

**DOI:** 10.1101/2021.05.04.21256604

**Authors:** Kevin Kahn, Richard M. Mariita

## Abstract

There is growing evidence that viruses responsible for pandemics, such as MERS and SARS, are mainly spread through aerosols. Recommendations have been introduced to reduce the transmission risk of virulent airborne viral particles by increasing ventilation rates, expressed in air-changes-per-hour (ACH), effectively improving the dilution of airborne pathogens via mechanical ventilation. However, the infrastructural and operational costs associated with upgrades of Heating, Ventilation, and Air Conditioning (HVAC) systems make these solutions expensive. It is well documented that UVC disinfection can help lower exposure risks by inactivating viruses in shared enclosed spaces, and the performances of such solutions be translated into equivalent ventilation (equivalent ACH or eACH). Here, we present the first framework to extract the optimum UVC output requirements for a target eACH, and improve facilities ability to comply with ventilation guidelines at lower energy costs. The Kahn-Mariita (KM) model considers the air quality of a shared enclosed space over time by supplementing existing mechanical ventilation with localized UVC air treatment, whether in recirculating units or upper-air systems, and extracts the systems requirements based on end-user needs by incorporating variables such as room size, occupancy, existing ventilation, and target eACH. An example of a conference room shows that a UVC chamber with an air recirculation rate of 160m^3^/h increases the ventilation from ACH=3 to eACH=7.9 and reduce down-time from 46 minutes to <10 minutes with as little 1W of UVC output. A recirculation rate of 30m^3^/h however offers no noticeable benefits above 200mW, with a maximum reachable eACH=3.9 and down-time of approximately 31 minutes. The KM model is unique in that it allows for the first time to find the optimum UVC output needs to ensure air quality is maintained and transmission risk minimized, while increasing energy savings. Recent studies suggest mechanically increasing fresh air supply will more than double the energy costs of HVAC systems, while the use of UVC reduces energy demand as much as by 50%. The KM model approaches air quality and energy efficiency in a unified way by incorporating UVC as a supplement to existing ventilation to increase eACH, reduce down-time, and increase the closed space occupancy.

## Introduction

The current covid19 crisis has highlighted the lack of readiness to managing pandemics globally and the need for solutions to treat the air, which is dominantly responsible for the transmission of pathogens [1] such as SARS-Cov-2 [2]. As temporary confinements implemented all over the world are slowly being lifted, there is an urgent need to implement safety measures to enable physical proximity between individuals, particularly in shared enclosed spaces such as offices, hospitals, schools, restaurants, and transportation systems [1]. Ultraviolet Subtype C (UVC) radiation, emitted at 200-280 nm with germicidal effect being able to disrupt nucleic acids (RNA or DNA) [3], can be used to disinfect air, thus managing disease outbreak [4]. UVC has exhibited effectiveness against bacterial hospital acquired infections [5], including the promise of lessening contact infections in a long-term acute care hospitals [6]. Additionally, studies carried out during the Covid-19 pandemic have demonstrated that UVC can rapidly inactivate SARS-CoV-2 [7] even at high viral titers [8]. Additionally, use of UVC to ensure clean air will save energy by ensuring reduced building HVAC energy demand [9].

With the world-wide accepted findings that airborne transmission route is the most dominant and highly virulent form of COVID-19 transmission [1] and with people gathering in closed spaces essentially share the same air [2], we have seen a global trend towards “healthy buildings”, driven by the urgency for automated, documented and reliable disinfection of air, surfaces and water in shared enclosed/inside spaces. Typically, air quality is ensured by ventilation, and increasing air-changes per hour (ACH), effectively diluting the virus by adding air and accelerating the reduction of airborne pathogen concentration [10]. Unfortunately, in most cases it is impractical to increase ACH because required improvements to existing HVAC systems is costly if at all possible with principal restrictions around ceiling heights, inability to upgrade the ducts or fan, and noise-air-flow-pressure drop imbalance due to non-optimal dimensioning that results from retrofit solutions. Additionally, with centralized air systems, the risks of cross-contamination, with air being re-circulated from one room to another remains [11]. In fact, except in hospitals, HVAC systems in closed public spaces are designed for comfort of occupants: odor control, removal of carbon dioxide, humidity and temperature control and may not have enough capacity for the right ACH for air-borne transmitted disease prevention [2]. It has been demonstrated that improperly applied HVAC systems may contribute to the transmission/spread of airborne diseases [12] as also suggested by descriptions from the Diamond Princess Cruise Ship [13]. This has also been indirectly proven using airflow-dynamics models [14] and experimental modeling [15]. Additional recement findings suggest that HVAC systems can facilitate SARS-CoV-2 transmission via shared air volumes with locations remote from areas where infected persons reside [11]. Moreover, HVAC contamination has been demonstrated in a Middle East Respiratory Syndrome (MERS) outbreak, caused by another Betacoronavirus [16].

Another complication for facilities management around increasing ventilation lies with the associated energy consumption from such solutions, as it is revealed by studies by Aviv et. al.,[9], Orme [17], Awbi [18], and Chenari et. al. [19]. New carbon-zero regulations, energy efficiency scoring, and credits create a strong incentive for managers of these assets to reduce ventilation, the primary source of energy consumption. Indeed, the higher the fresh-air injection rate, the more heating or cooling is required and the more expensive it is if systems are optimized with control to adjust with unpredictable changes in external temperature and humidity. For instance, rotary heat exchangers widely used in new constructions to save energy by capturing the heat have raised concerns due to potential transfer of viral load between the return and supply air flows. Conflicting reports on this account have pushed building managers to bypass them, highlighting the friction between energy efficiency and air quality.

In this paper, we present an alternative way of approaching the problem of air quality by using localized UVC for air treatment which is converted into an equivalent ventilation, which we will refer to eACH, (**Fig. 1)**: in other words, we look at quantifying the equivalent increase of mechanical ventilation to reach similar air quality within a defined amount of time. This allows to reach ventilation targets without upgrading existing HVAC system, and reversely provides an opportunity to optimize (reduce) existing ventilation already meeting recommended ACH by substituting with local UVC solutions. The Kahn-Mariita (KM) model presented here has the potential to rapidly allow buildings to comply with ventilation guidelines such as from ASHRAE [20] and WHO [21], to improve their energy efficiency rating, and provide a simple framework to reopening shared enclosed spaces, increase time spent in those shared enclosed spaces and manage occupancy limits, thus accelerating a return to “normal life”. The KM model is not limited specifically to SARS-CoV-2 and thus also offers a solution effective for any type of airborne microbial contaminants, such as the flu, tuberculosis or SARS-Cov-2, MERS etc.

**Fig. 1:**
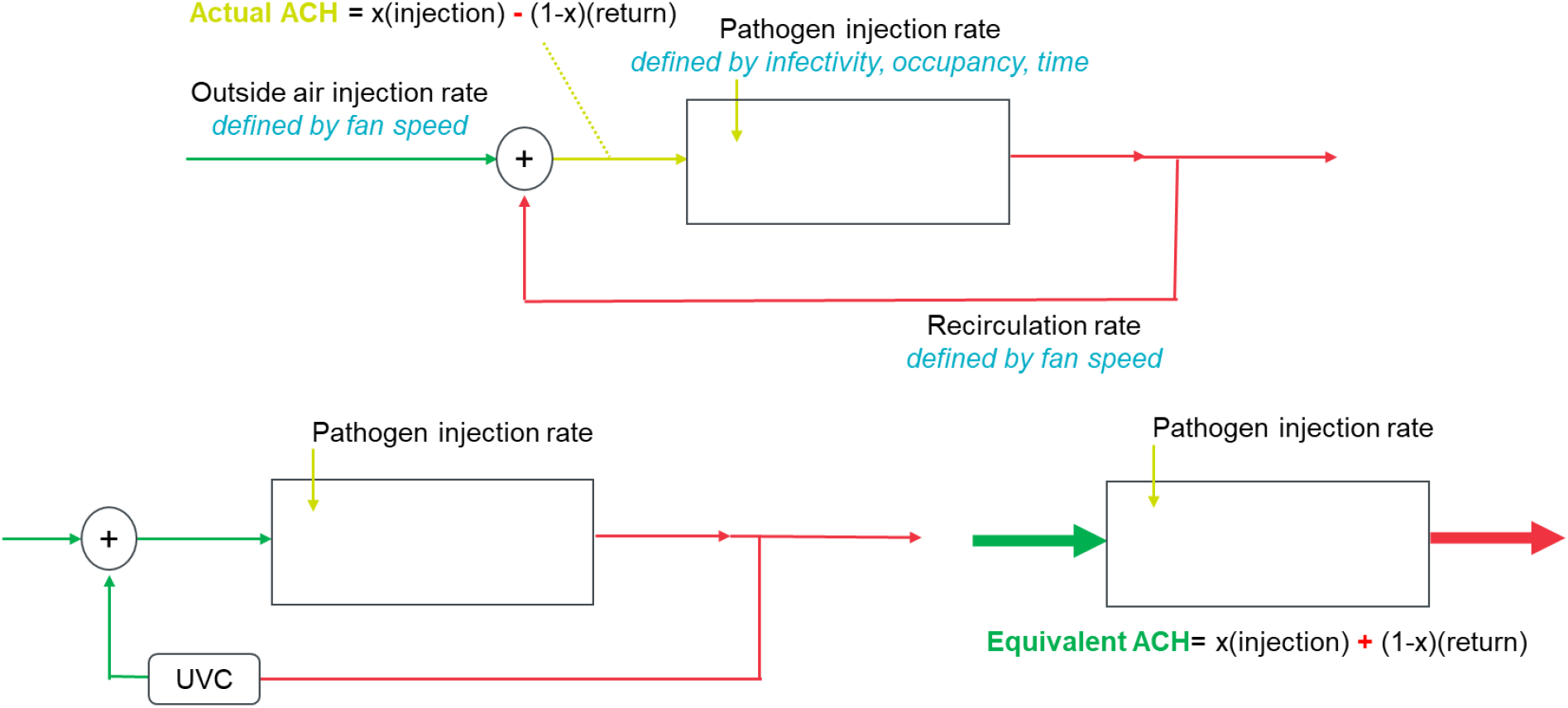
Equivalent ACH (eACH) as it relates to fresh-air injection rate

### The hypothesis-Airborne transmission of pathogens can be reduced and energy efficiency in shared enclosed spaces improved by adding UVC to re-circulating ventilation systems to obtain effective e-ACH target

The uniqueness of the KM model lies in the incorporation of UVC with existing ventilation. Previous studies from Walker and Ko [22] and Kane et. al., [23] have investigated the effect of UVC on airborne pathogen reduction and defined performances of UVC system independently of existing ACH. In particular, studies have linked i) UVC output to occupancy [24], which yields a 25mW/person requirement to increase equivalent fresh-air-injection rate by factor of 2, and UVC output to volume [25], which suggests 15-20mW/m^3^ requirement to reduce risk down to baseline risk. None of these studies look at the integrated performances with existing ventilation, nor do they allow to work back from a target ACH. Here, we try to provide a strong model to output a room or space eACH rather than a device performance, which allows for better facility management, and simplifying the integration of UVC systems for air treatment. The use of the KM model supports HVAC systems designed for closed shared spaces by helping understand the inactivation of viruses while achieving eACH, thus preventing the spread of viruses via HVAC systems as well as poorly ventilated shared enclosed spaces. This is even more important by recalling that some viruses in the Betacoronavirus genera such as SARS-CoV-2 are approximately 60-140 nm (0.06-0.14 micron) in diameter [26]. These viruses can easily pass through common filtration systems whose pore size is more than 1 micron since HEPA filters are only used in certain spaces such as isolation rooms or intensive care units (ICUs) [12]. The approach of e-ACH consists in assigning a quantitative measure, which consists of localized air treatment in poorly ventilated room with UVC light to reduce airborne contaminant concentration, which corresponds to the same air quality microbiologically to that of an increased mechanical ventilation. The model and methodology we present allow to quantify optical energy needs to achieve a defined log reduction, link it to a risk level and equivalent ventilation.

The KM model shows equivalent air-changes per hour by defining air quality level - 90% reduction of the steady state concentration without UVC-and by relating UVC solution to the amount of time saved to reach the same air quality, which utilizes the following steps:

1. Define the steady state concentrations of a system with existing ACH and no UVC applied. This refers to a number of infective patients that would enter the space and remain until the steady state concentration is reached. We are not interested in the time this process takes because it gives us the worst-case scenario. Any amount of time the patient would leave the room prior to that state reached would results in a lower concentration. A transient view shows this concentration is approached relatively quickly, typically within a minute.
2. Consider the infective person leaves the room, then we factor the speed of the exponential decay in the above concentration due to ventilation. This is solely determined by the ACH, which considers the volume of the room and air speed.
3. Add a UVC source within a recirculating chamber within the room. Here we model the efficiency of single-pass disinfection through the reactor design (size and materials UVC reflectivity) and the air flow which defers from the room ventilation and controlled locally. We compute the new steady-state concentration by adding the UVC on top of the existing ventilation, thus summing the clean air delivery rates.
4. Extract the ACH from the difference in steady-state concentration and input this into the decay model. This yields the time to reach the target concentration level.

The KM model microbial disinfection efficacy relies on UVC output, recirculation rate, existing ACH, and chamber design to obtain desired eACH. We find that the two most important factors are the existing ACH and recirculation rate, the first acting as a limiting element, and the second as the highest contributor to the increase in equivalent ventilation. UVC output affects the single-pass performance, with its additional relative contribution to eACH regressing. The increase in eACH in turn accelerates the time to reach a target concentration, which converges towards a minimum time.

The main problem with airborne pathogens is that in closed, poorly ventilated spaces the risk of transmission remains high for a long period of time [27]. Both calculations and experimental measures show aerosol droplets concentration is reduced by 63% [2] for every ACH as demonstrated by Rijn et. al.,[28], Nunayon et. al., [29] and by Bazant and Bush [30]. This means that in a typical office room with ACH=3, it takes > 46 min to reach a 90% pathogen concentration reduction. This view assumes no new source of contaminants or ignores the droplets that attach onto surfaces. Indeed, recent studies support the theoretical concepts presented here by showing how reduction in viral concentration affects the airborne transmission risk [31], and the results deviated from the linear relationship as the marginal benefit of incremental disinfection or log reduction value (LRV) is lower than the previous one. The key conclusions here are: i). There is little added value from aiming to higher concentration reduction values than 90% and ii). Because the risk is reduced the most by relatively low reduction rates, any recirculating system should be designed for higher air flow with low UVC dose rather than the conventional reactor designs traditionally used in water disinfection which aim at increasing the dose by reducing the flow. Here the dynamics is key, and recirculation needs to be considered, as well as how quickly the total air volume is treated.

For example, consider a room of 50m^3^, with an existing ACH of 3 (meaning it takes 46 minutes to reduce 90% concentration and bring transmission risk back to baseline risk), and a recirculating unit where air flows at two different velocities, 200m^3^/h and 20m^3^/h respectively, into a 70% UVC reflective chamber with 700mW of UVC optical output. Simple relationships allow to estimate the fluence rate from these parameters, which have proved to be a good prediction of microbial results. The higher power fan scenario “only” reduces 73% of pathogen in a single pass, while the lower speed fan which increases the exposure time to UVC of air leads to >99.9999% reduction (LRV6) per pass. It must be noted that in real term, to achieve such a high LRV value will require a very well-designed reactor since any deviation from all flow getting the exact same dose will have a large impact on the LRV (conversely, low LRVs are less sensitive to small variations). However, for the entire room to obtain a 90% reduction, there is acceleration from 46 minutes without UVC to 9 minutes for the higher fan power and to 35 minutes for the lower fan power, leading to eACH=8.5 and eACH=3.6 respectively. We therefore show that *dose or LRV is no longer the relevant metric for air treatment* -as it is in both water and surface disinfection where single pass matters and regulations aim for full reductions of specific pathogens. Instead the most important factors here are the existing ACH and the recirculation speed for the concentration to be sufficiently reduced to keep the risk close to the baseline. The eACH model captures these dynamics and incorporates variables such as volume of air to be treated, occupancy, reactor design, UVC output and wavelength, pathogen of interest, and only outputs the equivalent ventilation rate and improved time to 90% reduction (other notations used here: D90 or 1LRV).

### I) KM MODEL DYNAMICS

The airborne transmission risk *TR(t)* can be broadly defined as a sum of a baseline risk *R*_*β*_, which is a function of proximity between people *d*, and a time-dependent term which is a product of the risk associated with the pathogen concentration *R(C)*, and the exposure time *t* to the concentration *C* which will compound the risk. The time factor is modeled in a probabilistic manner with a location *μ* and scale *s* dependency.

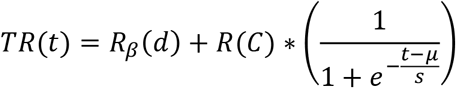

Ordinarily, in these applications, increasing the distance between people is the most efficient way to reduce contamination rate. The proximity effect is more pronounced when ACH is higher because the concentration reduces faster. On the other hand, as a person stays in a closed space, the time-dependent term (second term in Eq. 1) becomes the largest factor in the increase in airborne transmission risk. The goal of air disinfection is to bring that risk down converging towards the baseline risk *R*_*β*_(*d*).

#### 1) Concentration Build-Up

Kowalski defines the steady state concentration Css as the fraction of the contaminant release rate Rr and the clean air delivery rate (CADR), the product of the removal efficiency RE and the total air flow Q in the recirculating chamber [32].

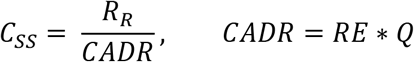

Css = steady state airborne concentration, cfu/m^3^

RR = source release rate, cfu/min

CADR = clean air delivery rate, cfm [32]

In the past, CADR has being used to assess the relative performance of filtration systems in recirculating chambers or that of UVC disinfection units [32]. While it is a good metric to compare devices, the question as to what is the required CADR or chamber performance to meet end-user needs is not accessible simply by looking into a chamber on its own. As we will show, this answer needs to incorporate the existing ventilation, the room volume and occupancy, and look at the marginal benefits from adding UVC as it relates to increasing occupancy, residence time and reducing down-time. The KM model therefore starts with CADR and links it to the end-user requirements. The advantage of CADR is the ability to measure it with microbial studies of single-pass performances, the risk however is to view this measurement as the final indication of performances.

A transient model allows for the estimation of the airborne concentration in a volume *V* as a function of time, and is defined as:

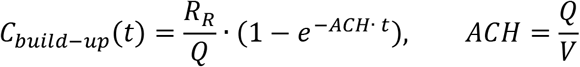

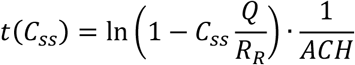

The relationship between ACH and LRV is easily extracted from the decay relationship, showing a concentration reduction per ACH RED% of 63.21% and an equivalent log reduction value of 0.43.

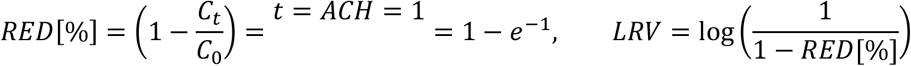

In our model, we first assume the initial concentration *C*_0_ = 0, and solve the equation over given amount of time at a defined contaminant release rate. The steady state for a system without UVC gives

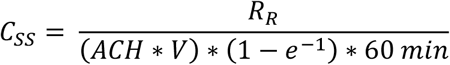

Previous studies have estimated the release rate for an infective person to be 99 RNA virus particles per minute, extrapolated from the weighted average of speaking and breathing particle release rate and the ratio of infective RNA containing virus particles to total released particles [33]. Susceptible subjects require a dose of 100-1000 RNA virus copies inhaled to cause an infection in 50% of susceptible [34]. The number of RNAs virus particles can be extracted from the volume of the particle in ml, and the concentration of RNA virus particles per exhaled particle of approximately 5. 10^8^ per ml [34]. Back to our example of a 50m^3^ room with an existing ACH=3 and one infective person, the equivalent average *C*_*ss*_ is 63 RNA virus particles/m^3^. It is straightforward to approach the transmission risk from the pathogen concentration factoring the breathing rate and time spent in the defined volume.

#### 2) Concentration Decay

The time for 1LRV (D90) to be reached is given by the decay equation where *C*_0_ = *C*_*SS*_:

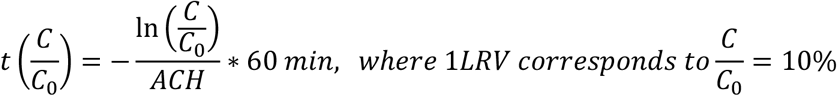

The time to 90% is independent of the room size, but the steady state concentration is. It is important to consider that the time to D90 is not a enough. Indeed, 90% of the steady-state concentration reached with ACH=2 is approximately the steady state concentration of ACH=20, thus if the goal is 90% reduction of a given steady-state concentration, it becomes a different target concentration ratio for the other ACHs. In this case, the time to reach the Css (ACH=2) with ACH=20 is 0, meaning it is already the permanent state reached by reducing the build-up process. This is exemplified by comparing the steady-state concentration at different ACHs, the time required to reach 90% reduction at the different levels and the time reduction from 90% reduction of a reference ACH (Table 1), where we show ACH=2 as reference. The benefits of increasing the ACH become very clear and the model helps quantifying the addition of UVC on the eACH and time reduction.

**Table 1.** Steady-state concentration at different ACH in a 50m3 room, and derived times to 90% reduction for each level and time to 90% reduction of a reference level - here ACH=2 - with increased ACH.

| **ACH** | **Css [RNA particles /m<sup>3</sup>]** | **Time to D90 [min]** | **Time to D90(Css(ACH=2))** |
| --- | --- | --- | --- |
| 2 | 93.97 | 69.08 | 69.08 |
| 3 | 62.65 | 46.05 | 28.17 |
| 4 | 46.98 | 34.54 | 18.94 |
| 5 | 37.59 | 27.63 | 13.49 |
| 10 | 18.79 | 13.82 | 3.56 |
| 15 | 12.53 | 9.21 | 0.94 |
| 20 | 9.40 | 6.91 | 0 |

#### 3) UVC Chamber: Concentration Build-up

Now, we add a UVC recirculating unit, which will increase the eACH in two processes: it will first lower the build-up concentration reached in the steady state model and will accelerate the decay time to the original D90 with a lower ratio required. First, we look at the steady-state concentration with UVC recirculation 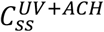, which is found by summing the CADR of mechanical ventilation (above) and that of UVC. The reduction factor RED depends on the UVC output and the design parameter of the UVC chamber (residence time, reflectivity). The relationships are shown in **Annex 1**.

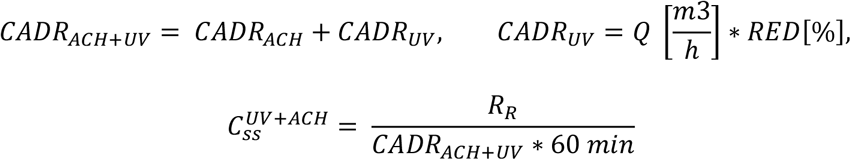

The build-up concentration can be understood as the risk for people present in the room and therefore defines the amount of time they can share the space. The decay concentration which we provide in the next and final step allows to plot the down-time for the risk to be minimized for a new person to enter the space once the infective subject has left the room. This saw-tooth wave function allows to consider risk in a dynamic mode, relevant to the different actors. Css can be calculated at various ACHs for a given system **(Table 2)**. It is evident from the calculations that the relative usefulness of UVC is higher at lower existing ACHs, and thus this understanding provides a framework for compliant buildings with high ventilation rates to reducing existing ventilation and maximize the use of UVC while reducing energy consumption. A win-win.

**Table 2.** Calculated Css at various ACHs for a 50m3 space, reactor volume of W*H*L (mm) of (200, 200, 400), a recirculating rate of 160m3/h, a target dose of 2mJ/cm2 (1LRV SARS-Cov-2, 265nm), and a UVC output of 500mW at 265nm

| ACH | $CADR_{ACH+UV}$ | $C_{ss}^{ACH}$ | $C_{ss}^{UV+ACH}$ | $C_{ss}$ Reduction |
| --- | --- | --- | --- | --- |
| 2 | 194.76 | 93.97 | 30.50 | 68% |
| 3 | 226.37 | 62.65 | 26.24 | 58% |
| 4 | 257.97 | 46.98 | 23.03 | 51% |
| 5 | 289.58 | 37.59 | 20.51 | 45% |
| 10 | 447.61 | 18.79 | 13.27 | 29% |
| 15 | 605.64 | 12.53 | 9.81 | 22% |
| 20 | 763.67 | 9.40 | 7.78 | 17% |

#### 4) UVC Chamber: Concentration Decay

The final step in the model considered considers that the steady-state concentration is reached and looks at the decay of the concentration assuming Rr=0 (infective person leaves the room for instance).

From the relationship below, we can extract the time required to reach the target concentration (90% of the non-UVC steady-state concentration 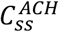) and the eACH.

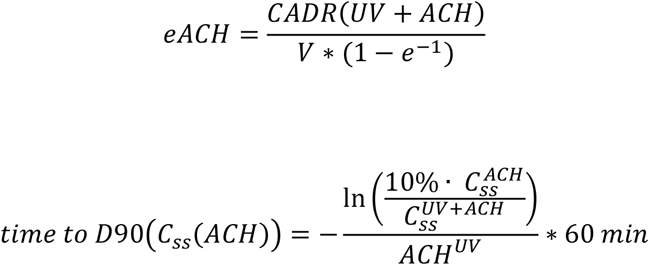

As the ratio of the target concentration level to the build-up concentration approaches 1 with higher ACH (remember the increase in ACH - or the eACH - reduces the build-up steady-state concentration), the time approaches 0. This can be well understood by the fact that the baseline risk remains dominant irrespective of the time spent in a space.

It is interesting to look at the effect of UVC output and recirculation rate on the e-ACH. First, let’s consider a change in output all else equal, with the parameters described above, and for an existing ACH=3, which provides the eACH reached with output ranging from 200mW to 10W of useful UVC output. It should be noted the LRV presented assumes that it follows a linear trend between dose and every additional LRV, which is not always the case. The model is abstracted of LRV of this problem since it considers the time to reach a single LRV (90% reduction), but it is useful to show that the relative benefits of UVC dose on the eACH decrease with increased dose. There is no noticeable difference in eACH or time reduction beyond 1W (eACH 7.9 and 9min to D90) and the best use of energy around 500mW (eACH 7.2 and 11min to D90), compared to the 46min without UVC (Table 3).

**Table 3.**
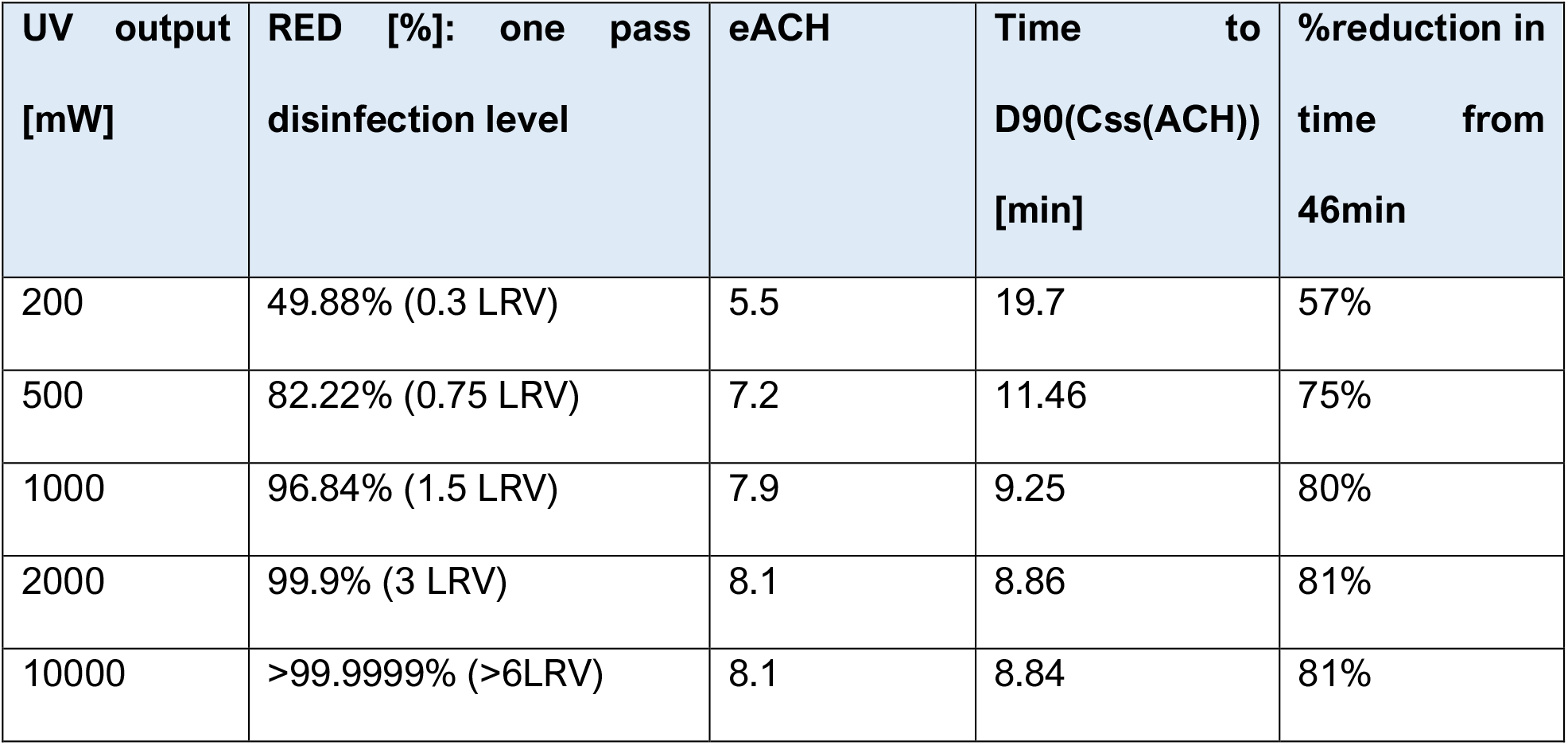
Effect of UVC on the eACH and time to D90(Css(ACH)) for a 50m3 room with existing ACH=3. The UV chamber parameters are 160m3/h recirculation rate, 70% internal reflectivity and wavelengths are normalized to 265nm

| UV output<br>[mW] | RED [%]: one pass<br>disinfection level | eACH | Time to<br>D90(Css(ACH))<br>[min] | %reduction in<br>time from<br>46min |
| --- | --- | --- | --- | --- |
| 200 | 49.88% (0.3 LRV) | 5.5 | 19.7 | 57% |
| 500 | 82.22% (0.75 LRV) | 7.2 | 11.46 | 75% |
| 1000 | 96.84% (1.5 LRV) | 7.9 | 9.25 | 80% |
| 2000 | 99.9% (3 LRV) | 8.1 | 8.86 | 81% |
| 10000 | >99.9999% (>6LRV) | 8.1 | 8.84 | 81% |

It is evident that the principal factor in increasing the ACH is the recirculation rate, that is if too low no matter how much output, only a fraction of the volume will be treated within the time for 1LRV to occur “naturally” with existing ventilation. We show this by comparing two sets of data (**Table 3** and **Table 4)**, which look at the same system except the recirculation rate is reduced from 160m3/h to 30m3/h. The single-pass disinfection rate rises of course but the maximum eACH reachable and disinfection time is reduced dramatically. Additionally, there is almost no difference between 200mW and 10W (50x higher outptut) on the eACH (Table 4), even though the disinfection per pass is increased by a couple of orders. The key is therefore to understand the limiting factor in the system, defined as the parameter which prevents the increase in other variables to translate into increase in eACH. In this example, it is the recirculation rate.

**Table 4.**
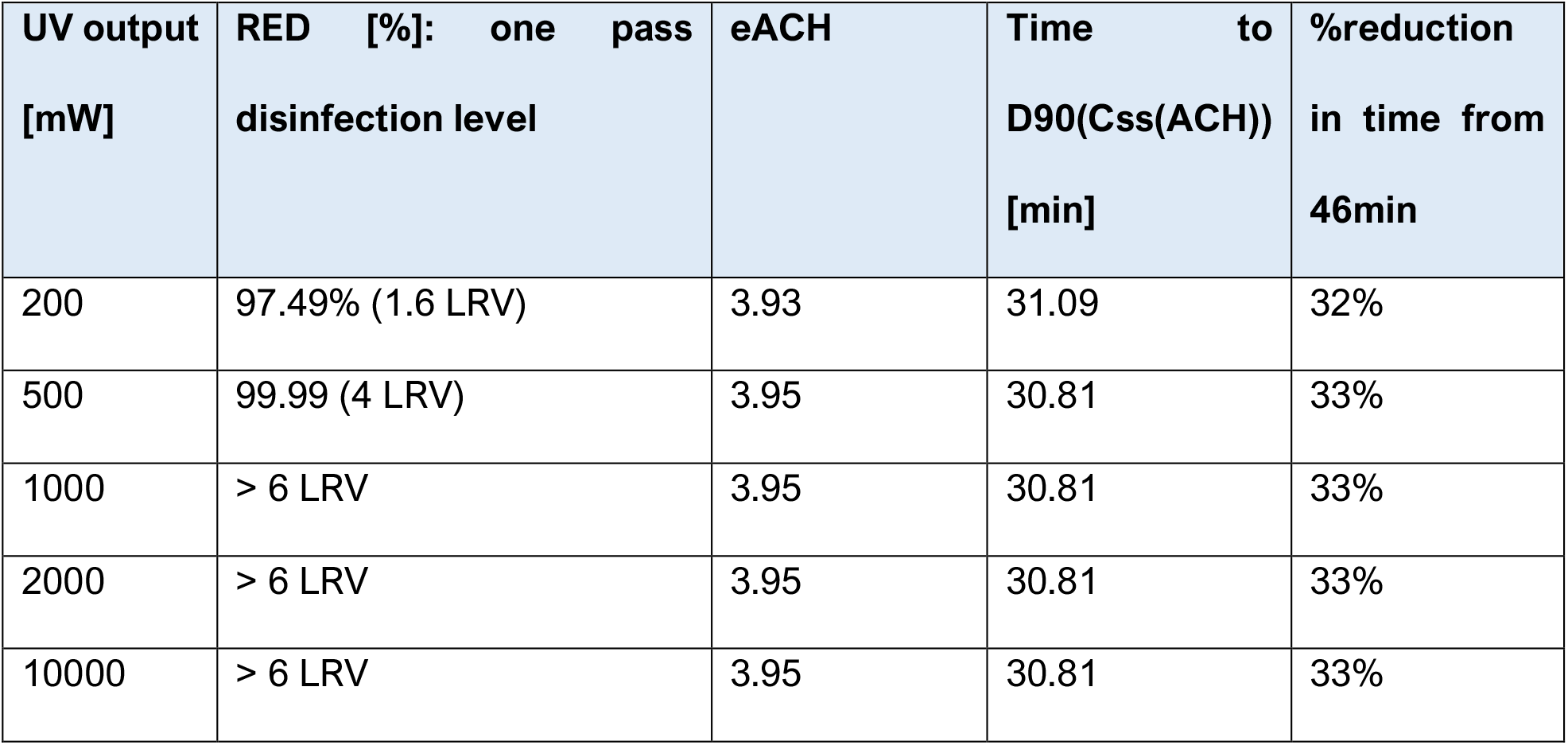
Impact of UVC on a similar system presented in Table 3, however with recirculation rate reduced from 160m3/h to 30m3/h

| UV output<br>[mW] | RED [%]: one pass<br>disinfection level | eACH | Time to<br>D90(Css(ACH))<br>[min] | %reduction<br>in time from<br>46min |
| --- | --- | --- | --- | --- |
| 200 | 97.49% (1.6 LRV) | 3.93 | 31.09 | 32% |
| 500 | 99.99 (4 LRV) | 3.95 | 30.81 | 33% |
| 1000 | > 6 LRV | 3.95 | 30.81 | 33% |
| 2000 | > 6 LRV | 3.95 | 30.81 | 33% |
| 10000 | > 6 LRV | 3.95 | 30.81 | 33% |

At higher recirculation rate, output matters more, however we see again a non-linear contribution between UVC output and its relative increase in eACH or decrease in time to D90(Css(ACH)). At 160m^3^/h, we find that 500mW provides 93% of the benefits of 10W. It must be highlighted that one important assumption in the model does not consider re-mixing after every pass in the chamber.

##### Upper air systems

While we have shown the limiting factor to be the recirculation rate in air chambers, upper-air systems would of course not be subject to this limitation. Instead, the KM model can be adopted for upper-air to use the existing ACH as recirculation rate, assuming perfect and constant air mixing (which is typically one of the main assumptions in epidemiological models such as Wells-Riley) [30], as well as a “UVC corridor” acting as a chamber with 0% reflection, meaning the UVC is fully absorbed by either the ceiling of the opposite wall.

Another important concept of the KM model is the fact that UVC is seen as a complementary addition to existing ventilation, therefore offering an integrated model of actual performances. A recent study using upper-air systems with both low-pressure mercury lamps and UVC LEDs as light sources showed a reduction time to 90% of initial concentration is just under 2minutes from 14minutes naturally was achieved with both sources [29]. The performances of the 200mW LED powered upper-air system were similar to a 21W lamp one, which is explained by the fact that most of the lamps output remains in the upper-air module: the actual UVC output which is transmitted out of the upper-air module is estimated to be around 375mW considering the conversation electrical-optical energy (33%), the normalization to germicidal power at 265nm (90%), and the systems loss due to the loovers (6% efficiency). The KM model predicts that the 200mW system should have accelerated the risk reduction to 4.2min and the 375mW (lamp) to 3.4min yielding an eACH of 22 and 25 respectively. While the overall performances are well captured, we attribute the differences from the air recirculation in the room which is not properly included. In fact, upper-air experiments include the use of fans for air recirculation, which is not included here and would improve the performances as we’ve shown the importance of recirculation.

### II) QUANTIFYING BENEFITS

While the KM model allows for output eACH, its extracted time to D90 (equivalent to a potential definition of down-time for a shared indoor space before re-use), the benefits of eACH is readily found by inputting the new-ACH into the model from Bazant and Bush from MIT [30] to quantify the benefits to end-users in terms of additional number of people or time allowed into a shared enclosed space thanks to the improved air quality level. For a volume of 50m3 (a typical conference room of app. 4×5×2.5 m^3^), the proximity rule of two-meter between every occupant would limit the occupancy to 6 people. With ACH=3, a maximum of 3 people are allowed without mask for 1hour if infective person is in the room to limit transmission risk. With ACH=12, this number is increased to 7. ^*^The guideline restricts the probability of airborne transmissions per infected person to be less than the risk tolerance over the cumulative exposure time listed.

The model developed by Bazant and Bush [30] allows to find the maximum occupancy at different ventilation rates, which in turn allows for a direct comparison of the three approaches, namely the UVC/occupancy, UVC/volume, and the KM model on equivalent ventilation is presented here. For a 50m3 room a maximum of 5 people are allowed at an ACH=7, with the eACH being reachable at various outputs (Table 3). The eACH=8 is the maximum reachable but does not allow additional benefit in occupancy from eACH=7). With a minimum output requirement of 25mW/person, the first approach yields a 125mW requirement for highly reflective chambers (PTFE >90%). This number would be directly affected by the UVC reflectivity of the material used in these designs, more so than the other approaches. For instance, an aluminum design (UVR 70%) would increase this value to approximately 400mW. The second approach on volume of 15-20mW/m3 yields 750-1000mW requirement. The difference between the two can be understood from the occupancy limitation imposed on the first, and thus the higher requirement considers a maximum fill up of the space. None of these considers the existing ventilation, nor the number of infective persons in the room. The KM model does and outputs for a target eACH=7 from an existing ACH=3 with a requirement of 500mW at 160m3/h recirculating rate (Table 5). The difference from the occupancy model is that as it increases, the risk shifts from concentration in the air to the proximity, which is well captured: the KM proximity provides an output equivalent to the occupancy up the point where proximity restrictions are broken down. The volumetric and occupancy approaches converge as occupancy is increased to fill the volume, however the hypothesis given here is that increasing output will not affect the risk level beyond a certain point. These models are also independent of the number of infective persons, while our model allows to incorporate this in larger spaces where the probability of multiple infection sources is higher than in the smaller spaces presented in these examples.

**Table 5.** Recommended UVC output by the three approaches (UVC/occupancy, UVC/volume, and UVC/target eACH)

|  | Output/Occupancy | Output/Volume | KM Model for<br>UV/target eACH |
| --- | --- | --- | --- |
| **Recommended**<br>**output** | 125 - 400 mW | 750 - 1000 mW | 500 mW |

## Conclusion

It is challenging to eliminate air-recirculation such that only fresh air is supplied to occupants in shared enclosed spaces. The KM model offers blueprints that reduce airborne transmission of pathogens by adding UVC to localized systems and converting the benefits into an equivalent air-changes-per-hour, thus complementing existing ventilation. The model particularly allows to walk back from a target eACH given by benefits such as increase in occupancy or time spent in shared enclosed spaces and output a UVC requirement as a function of UVC, space volume, existing ventilation, recirculation rate and single-pass performance. This allows for devices to be tested on their single-pass performance, and link to the eACH as a function of the recirculation rate and the existing ventilation. Finally, and of importance for buildings sustainability, the KM model allows to balance centralized ventilation with localized UVC air treatment options to decrease dependency on outside air injection and increase energy efficiency. This unified view of air quality and energy also provides an alternative approach to upgrading buildings to meet new carbon zero requirements.

## Data Availability

The authors confirm that the data supporting the findings of this study are available within the article and its supplementary materials.

## Conflicts of interest

Both K.K. and R.M.M. work for Crystal IS, an Asahi Kasei company that manufactures UVC LEDs.

## Funding information

This work received no specific grant from any funding agency

## Author Contributions

**Kevin Kahn**: Implemented the idea for this model, contributed towards the literature search, drafted the initial manuscript and revised the final manuscript;

**Richard M. Mariita**: Contributed towards review of model, literature search, contributed to writing of initial manuscript and revised the final manuscript.

## Acknowledgements

We are grateful to Crystal IS Inc. and Asahi Kasei for their support, particularly to Eoin Connolly and Amy Miller. Authors sincerely thank Prof. Leo Schowalter and Dr. Sébastien Blumenstein for their useful insights and for proof-reading the manuscript, and Pasquale Attrotto for his valuable contribution, including suggesting and validating some relevant formulae used to describe phenomena presented in this work.

## Consent Statement/Ethical Approval

Not Required

## About the Authors

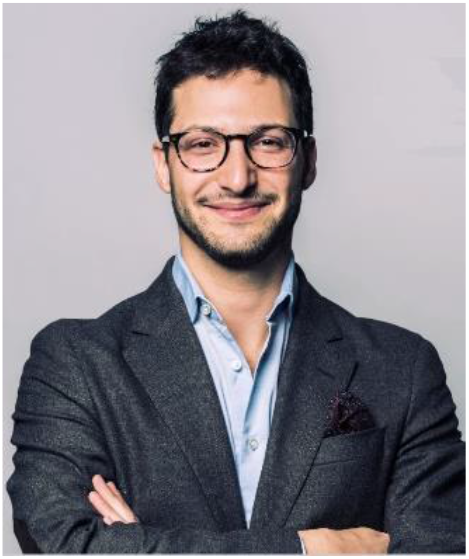

**Dr. Kevin Kahn** was born in Switzerland and received a BSc in electrical engineering from EPFL, an MRes and PhD in applied physics from Imperial College London, and an MBA from the University of Chicago Booth School of Business. He was a research fellow at the National University of Singapore, where his work focused on III-nitrides and two-dimensional semiconductors. Dr. Kahn has since been working at Crystal IS, an Asahi Kasei company, specializing in UVC LED solutions for infection prevention, and currently leads market development for EMEA. He was elected to the technical committees of Watercoolers Europe, Lighting Europe, vice-chair for IEEE standard on air treatment, and his work has been awarded the 2018-2019 IUVA best paper award.

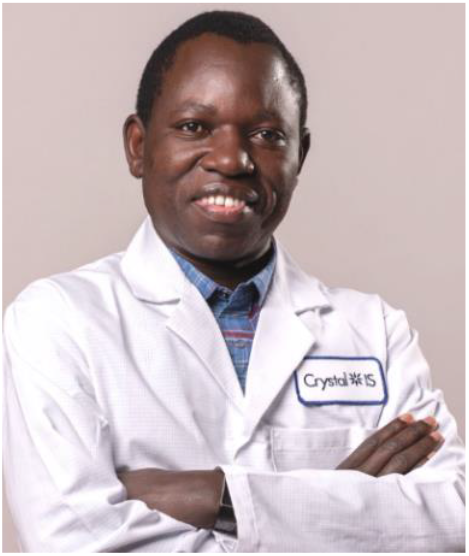

**Dr. Richard M. Mariita** was born in Kenya and completed his MSc. in Medical Microbiology from Kenyatta University while performing tuberculosis research from a CDC affiliated BSL-3 Lab at the Kenya Medical Research Institute (KEMRI). Upon completion of his MSc., he came to the United States to pursue his Ph.D. and Postdoc. Dr. Mariita was a visiting Scientist at Brown University. Dr. Mariita is recipient of numerous awards, including: Brown University’s Presidential post-doctoral fellowship, College of Sciences and Mathematics Dean’s best research award (Auburn University), The Joseph Kirby Farrington endowed Fund award for excellence towards his exceptional achievement, Selman A. Waksman Endowed Scholarship in Microbial Diversity at the MBL in Woods Hole, MA, Bernard Davis Endowed Scholarship Fund at the MBL in Woods Hole, MA and was Auburn University’s Cell and Molecular Biology Ph.D. fellowship winner. Currently, Dr. Mariita is a Senior Microbiologist at Crystal IS- an industry-leading manufacturer of high performance UVC LEDs. He leads microbial disinfection performance investigations of UVC LED prototypes and products. Through the BSL-2 lab at Crystal IS, Dr. Mariita and his team are at the forefront of using microbiological expertise to validate existing products as well as help improve prototypes with the sole goal of coming up with new products that offer the desired air, water, and surface disinfection solutions to customers. Dr. Mariita is a committee member of NSF/ANSI 55 as well as a member of IEEE working group whose goal is to come up with a standard on air treatment. He is also a member of American Society for Microbiology (ASM) and Microbiology Society (Europe).

## ANNEX1

Supporting information on computing the LRV in a UVC chamber

The dose associated with a log reduction value (LRV) for a defined pathogen reached in a single-pass UVC chamber is estimated under an assumption of linearity, using the following relationships.

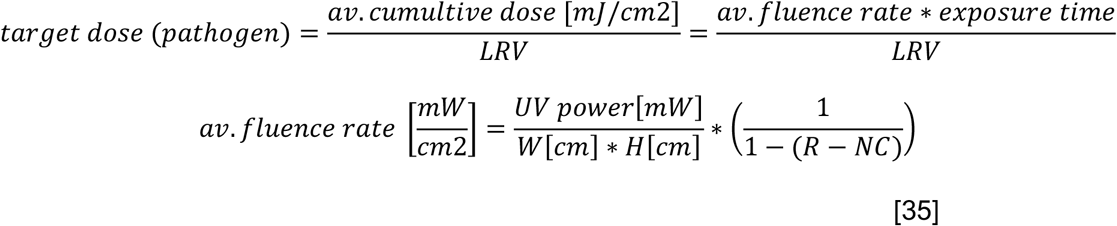

The fluence rate is Where *R* is the reflectivity in % and *NC* the ratio of surface area not covered by reflective material (for instance if an LED strip is fixed on top of a reflective surface or end caps of the chamber are used for the fans).

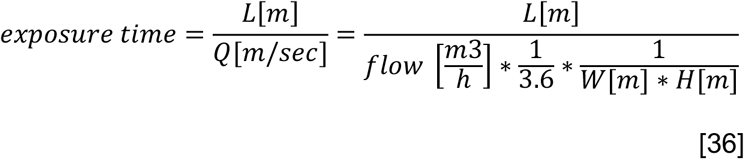

## Notes

### Competing Interest Statement

The authors have declared no competing interest.

### Funding Statement

No external funding was received

### Author Declarations

No IRB/oversight body was required

